# Dyslipidemia is a metabolic hallmark of acute pain in sickle cell disease

**DOI:** 10.64898/2026.06.24.26356495

**Authors:** Jonathan D. Enders, Zainab Khalid, Vivien Blecking, Allison D. Ebert, Amanda M. Brandow, Cheryl L. Stucky

## Abstract

Individuals with sickle cell disease (SCD) experience intense acute episodic pain associated with vaso-occlusive events and persistent, often daily, chronic pain. Triggers for acute episodic pain include cold exposure, strenuous exercise, and hypoxia. The molecular mechanisms underlying acute pain in SCD are poorly defined. We asked whether acute pain was associated with an altered metabolomic profile in individuals with SCD. We performed untargeted metabolomics on plasma from 25 children with SCD obtained during two disease states: 1) during an acute pain episode, and 2) during baseline state of health (“baseline health”). Control plasma was analyzed from 25 race-matched healthy controls. We identified 318 dysregulated metabolites in SCD patients during baseline health relative to healthy controls. Baseline health SCD samples had elevated pyrimidine, polyamine, and methionine metabolites, whereas arginine and sphingomyelin metabolites were decreased. During acute pain, we identified 448 dysregulated metabolites relative to baseline health conditions in the same SCD patients. We found decreased amino acid metabolites and acyl-carnitines, consistent with hypoxia. Network analysis revealed eight metabolic modules that were significantly differentially correlated to healthy controls, baseline health, or SCD acute pain. Modules enriched for porphyrin metabolism were correlated with SCD during acute and baseline health conditions. Other modules identified prominent dyslipidemia during acute pain in SCD relative to baseline health and healthy controls. Furthermore, we identified a metabolic module characterized by multiple sphingomyelins that were reduced in SCD and correlated with acute pain. Our findings identify dyslipidemia and impaired oxidative metabolism as potential drivers of acute pain in SCD.

## Introduction

Sickle cell disease (SCD) is the most common inherited hemoglobinopathy, affecting nearly 8 million individuals globally^1–3^. SCD arises from mutations in the β-hemoglobin gene, rendering hemoglobin susceptible to misfolding and aggregation, leading to sickling of red blood cells (RBCs), disruption of microvascular function, and occurrence of localized vaso-occlusive events. Indeed, these vaso-occlusive events are the most common complication of SCD, leading to abrupt onset of intense acute pain that can last for days in affected adult and pediatric populations^4–6^. These acute pain events in individuals with SCD are often refractory to conventional analgesics^7^. Recurrent pain episodes increase in frequency and severity as individuals with SCD age into adolescence and adulthood, and a higher rate of pain is associated with mortality in adults. Acute pain, therefore, has a substantial impact on an individual’s health-related quality of life. This morbidity and increased mortality associated with acute pain underscores the importance of studying this common complication of SCD.

The pathophysiology of acute pain in SCD remains poorly defined. Vaso-occlusive events in SCD are triggered by exposure to cold, exercise, and hypoxia^6^, each of which is associated with compensatory metabolic adaptations. Hypoxia is of particular interest given its association with ischemia-reperfusion injury that occurs in SCD and downstream complications, such as inflammation. Metabolic adaptations related to hypoxia include reduced flux through the tricarboxylic acid (TCA) cycle, reduced mitochondrial oxidative metabolism, and increased reactive oxygen species (ROS) generation^8^. Amino acid uptake and utilization are likewise affected by hypoxia^9–11^, and negatively affect TCA metabolite availability, anabolic processes, and antioxidant defense. Prior work has established that individuals with SCD have reduced amino acids during baseline health and acute pain presentation^12,13^. However, these studies were either unable to conduct direct pairwise comparisons within the same individual between disease states or relied on targeted metabolomic approaches^14–16^, potentially missing important and related metabolic pathways that are altered during acute pain in SCD.

To capture a holistic snapshot of metabolic adaptations during baseline health and acute pain presentation in SCD, we performed untargeted metabolomics on plasma collected from 25 children with SCD during clinical presentation with acute pain and during baseline state of health. These samples were further compared to healthy race-matched controls. Our results revealed dysregulation of arginine and methionine pathways during baseline health and exacerbation of these pathways during acute pain. We also found changes in bioenergetic fuel availability consistent with acute hypoxia. Network analysis further revealed that individuals with SCD have dyslipidemia, which is associated with both acute pain presentation and the number of acute care visits in the emergency department or hospital admission. Together, our findings reveal a unique metabolic fingerprint in children living with SCD and identify novel targets for metabolic intervention to alleviate acute pain in SCD.

## Methods and Materials

### Study Population and Clinical Data

Plasma from individuals with SCD and healthy controls was collected under an institutional human research review board-approved protocol (MCW IRB PRO00047854). Written informed consent/assent was obtained from the study participant and/or legal guardian as appropriate. Plasma samples were collected from individuals with SCD (7-19 years of age) during hospitalization for acute pain (SCD Acute) and baseline state of health during an outpatient clinic visit (SCD BL). Baseline state of health was defined by the absence of acute care visit for pain or other SCD complication, including infection, within the preceding two weeks prior to plasma collection. Individuals with SCD on chronic transfusion therapy were excluded. Plasma samples were collected from age-and race-matched healthy individuals without SCD (HC). Diagnoses with other pain-related or neurological diseases were used as exclusion criteria (SCD and controls). Demographic and disease characteristics were collected from the medical record. Total number of acute care encounters for pain within the 5 years prior to plasma collection were abstracted from the medical record. These included emergency department treat and discharge visits and hospitalizations for pain.

### Human plasma metabolomics

Samples from individuals with SCD (Acute and Bsl) and healthy controls were collected through routine venipuncture, processed, and immediately stored at-80 °C until use. SCD Acute samples, paired SCD baseline samples, and HC samples (n=25/group) were analyzed by Metabolon, Inc., using in-house methods for sample processing, UHP/LC tandem-mass spectroscopy, and metabolite peak identification, quantification, and normalization.

### Comparison of metabolomics between groups

Concentrations of individual metabolites were compared using Welch’s 2-sample t-test (SCD Acute vs HC and SCD BL vs HC) or paired t-test (SCD Acute compared to SCD BL) to provide a raw p-value. False discovery rate (q-value) was calculated using the Storey-Tibshirani method^17^. Metabolites with a q-value less than 0.1 were considered significantly different between specified comparisons.

### Metabolite Set Enrichment Analysis (MSEA)

For downstream MSEA, metabolites with a p-value of less than 0.05 were considered significantly dysregulated. Lists were generated containing all significantly upregulated and significantly downregulated metabolites across each comparison. MSEA was then performed for each of these lists using the Small Molecule Pathway Database (SMPDB) metabolite set library in MetaboAnalyst 6.0^18^.

### Weighted Gene Correlation Network Analysis (WGCNA)

Network analysis was performed on normalized metabolite data from Metabolon using the *WGCNA* package in R^19^. Data were filtered to exclude metabolites of unknown structure or those that were not present in all samples, resulting in 953 metabolites for analysis. Soft-thresholding power was set to the lowest power achieving a scale-free topology fit of R^2^>0.8. An unsigned network was built using a topological overlap matrix, and modules were detected using hierarchical clustering and dynamic tree cutting with a minimum module size of 30 metabolites. Module eigengenes were correlated to sample group and acute care utilization data (emergency department visits, inpatient hospitalizations, and total care visits). A correlation coefficient of +/-0.25 and a significant correlation (p < 0.05) was used to identify significant module eigengene correlations. Hub metabolites were identified using eigengene-based connectivity (kME).

## Statistical Analyses

All statistical analyses were performed using R (version 4.5.2) and RStudio. Data were analyzed by two-tailed t-test (SCD BL vs HC) or paired t-test (SCD Acute vs SCD BL). False discovery rate was determined using the Storey-Tibshirani method^17^. Data were considered statistically significant with a p-value of less than 0.05 and a q-value of less than 0.1. Confidence intervals are presented as the standard error of the mean.

## Results

### Study Population

We performed unbiased global metabolomics on plasma isolated from twenty-five children and adolescents with SCD during acute pain events and at baseline health. We additionally recruited twenty-five race-matched healthy controls. Demographics of our study population are presented in **Table 1**. There were no differences in sex between individuals with SCD and controls; however, the mean age of individuals with SCD was significantly lower than that of healthy controls.

**Table 1.** Patient demographics.

Figures
| **Table 1. Patient demographics** |  |  |  |
| --- | --- | --- | --- |
|  | <u>Healthy Control</u> | <u>Sickle Cell</u> | <u>p-val</u> |
| Sex |  |  |  |
| Female | 13 (52%) | 14 (56%) | 0.99 |
| Age (years, 95% C.I.) | 12.8 (11.7-13.9) | 10.8 (9.3-12.2) | 0.022 |
| Race |  |  |  |
| African American | 100% | 100% | 1.0 |
| Genotype |  |  |  |
| SS |  | 17 (68%) |  |
| SC |  | 6 (24%) |  |
| SB Thal |  | 1 (4%) |  |
| S-Oarab |  | 1 (4%) |  |
| Number ED Visits (*range*) |  | 4 (0-25) |  |
| Number Inpatient Visits (*range*) |  | 10.5 (0-29) |  |

### Individuals with Sickle Cell Disease (SCD) exhibit unique metabolomic signatures at baseline health and during acute pain events

Unbiased metabolomics identified 318 unique compounds that were significantly up-or downregulated between SCD BL and healthy controls (**Figure 1A**; **S. Table 1**) and 448 significantly up-or downregulated compounds between SCD Acute and SCD BL (**Figure 1B**). We performed dimensionality reduction with principal component analysis (PCA) to visualize differences due to metabolites and observed separation between healthy controls, SCD BL, and SCD Acute groups (**Figure 1C**), indicating distinct metabolic profiles between individuals with SCD during acute pain episodes, individuals with SCD during baseline health, and healthy controls. Further analysis revealed significantly increased products from metabolism of analgesics in the SCD Acute samples relative to healthy controls (**Figure 1D**). This finding is consistent with the treatment of pain in the SCD Acute group but may represent an artificial driver of separation between SCD Acute and SCD BL samples. Therefore, we performed an additional PCA on metabolomic data excluding xenobiotics (**Figure 1E**). Visualization of these principal components also demonstrated separation between the healthy control, SCD BL, and SCD Acute groups, indicating that differences between the biochemical profiles of SCD Acute, SCD BL, and healthy controls are not simply driven by use of analgesics.

**Figure 1.**
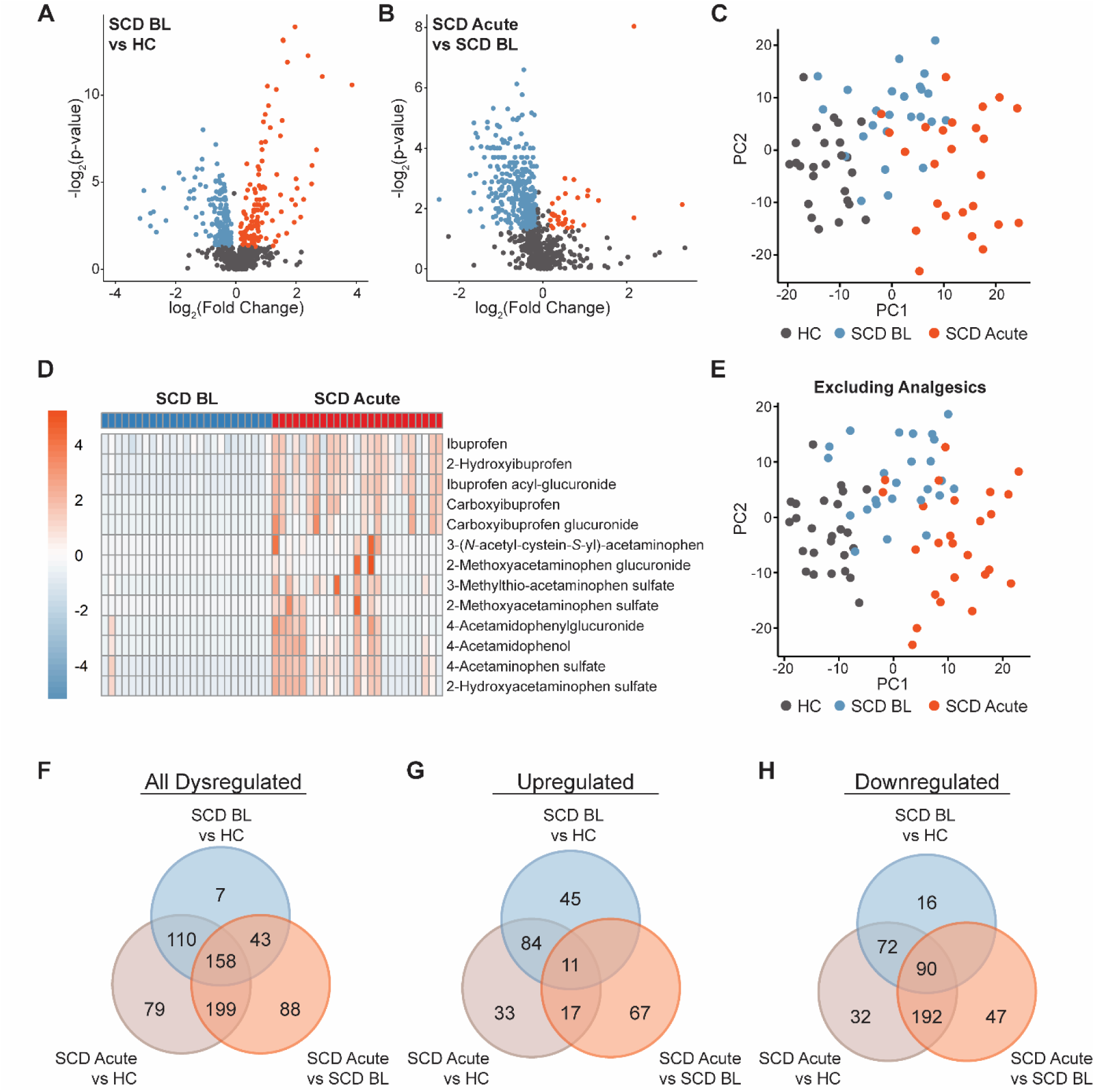
Children and adolescnets with sickle cell disease (SCD) exhibit distinct metabolic signatures during baseline health and acute pain. Untargeted metabolomics on plasma identified 318 differentially regulated metabolites between children and adolescents with SCD at baseline health (SCD BL) compared to healthy controls (HC, *A*) and 448 differentially regulated metabolites between SCD acute pain (SCD Acute) and SCD BL groups (*B*). Separation between HC, SCD BL, and SCD Acute metabolic profiles was visualized after dimensionality reduction by principal components analysis (PCA, *C*). Analgesics and metabolites of analgesic catabolism were elevated during SCD acute pain relative to baseline health (*D*). Visualization of HC, SCD BL, and SCD Acute metabolic profiles shows separation after removing xenobiotic metabolites from PCA (*E*). Overlap between all dysregulated metabolites (*F*), only upregulated metabolites (*G*), and only downregulated metabolites (*H*) for SCD BL and HC, SCD Acute and HC, and SCD Acute and SCD BL comparisons.

We reasoned that the presence of metabolites from analgesics may obscure downstream analyses of metabolic pathways by metabolite set enrichment analysis (MSEA). Therefore, we omitted xenobiotic metabolites from pharmaceuticals and dietary consumption to generate lists of metabolites that were significantly increased or decreased (**Figure 1F**), only significantly increased (**Figure 1G**), and only significantly decreased (**Figure 1H**) across comparisons.

### Reduced arginine metabolites are associated with increased polyamine, methionine, and pyrimidine metabolism during baseline health in individuals with SCD

Significantly increased and decreased metabolites in individuals with SCD at baseline health compared to healthy controls are presented in **S. Table 1**. Compared with changes in individual metabolites, metabolite sets provide better insight into biological and physiological adaptation. MSEA of upregulated and downregulated metabolites in individuals with SCD at baseline health compared with healthy controls revealed increased metabolism of polyamines, bile acids, pyrimidines, and methionine (**Figure 2A**) and diminished arginine metabolism (**Figure 2B**). Arginine and methionine metabolism converge during polyamine metabolism (**Figure 2C**). Our observed decrease in arginine metabolism is driven by greater reductions in ornithine than urea cycle metabolites (**Figure 2D**), suggesting that elevated polyamine biosynthesis (**Figure 2E**) is likely due to increased flux from arginine during baseline health in SCD. Although methionine metabolism is enriched among upregulated metabolites, methionine metabolites that are required for glutathione biosynthesis were decreased during SCD baseline health (**Figure 2F**). In addition to its roles in protein synthesis and glutathione biogenesis, methionine supports the folate cycle, in turn contributing to nucleotide synthesis. Nearly all metabolites associated with pyrimidine metabolism detected in our analysis were elevated in SCD at baseline health (**Figure 2G**). Thus, differential utilization of arginine and methionine in biosynthetic processes appears as a major component of metabolism in SCD during baseline health.

**Figure 2.**
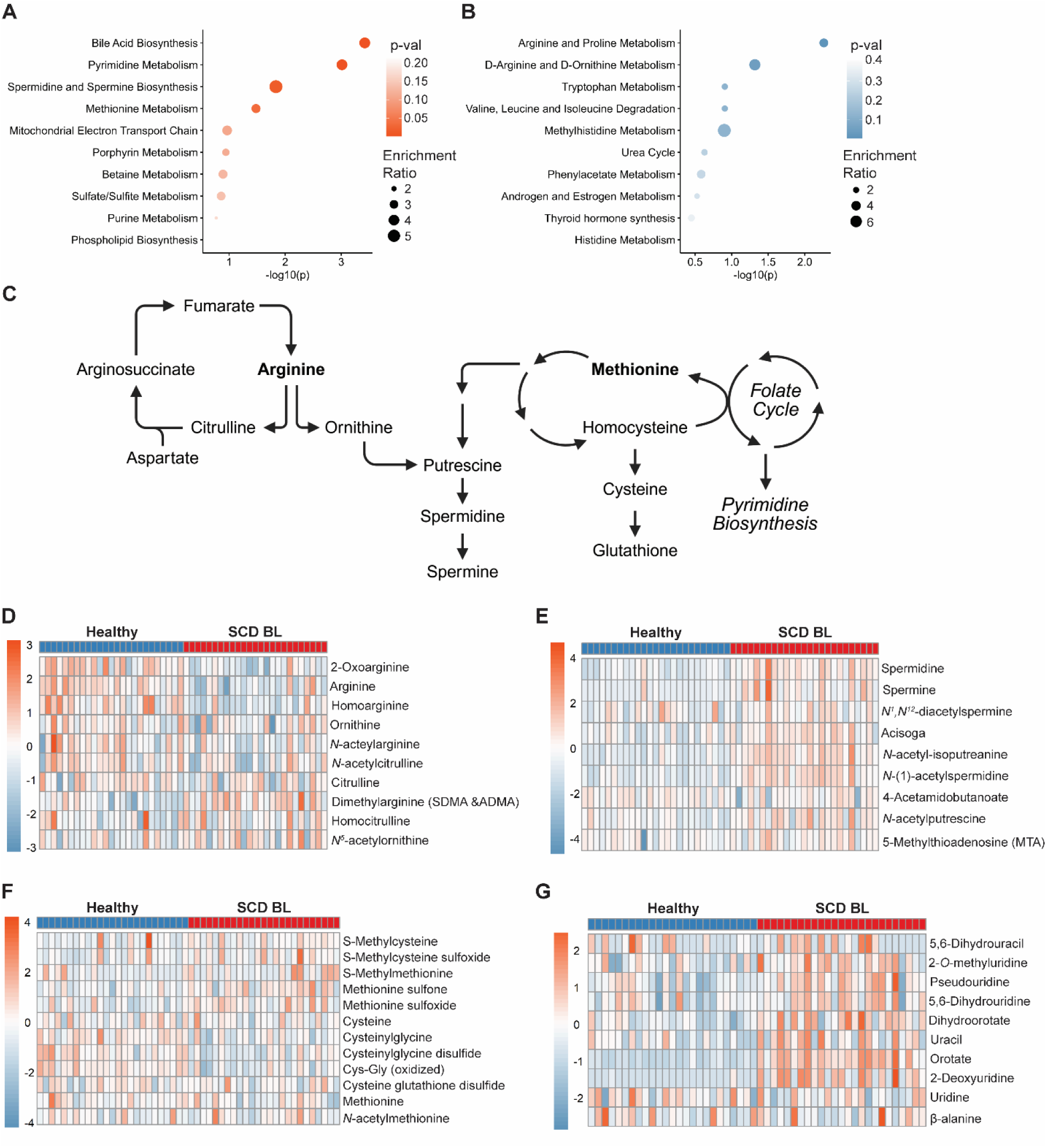
Arginine deficiency is associated with elevated polyamines, methionine, and nucleic acid synthesis during SCD baseline health. (*A*) Metabolite set enrichment analysis (MSEA) of upregulated metabolites during SCD baseline health reveals elevated bile acid, polyamide, and pyrimidine synthesis and methionine metabolism. (*B*) Downregulated metabolites during SCD baseline health are enriched for pathways associated with arginine. (*C*) Arginine and methionine metabolism converge at polyamine synthesis. Individual metabolites associated with arginine are diminished in SCD BL (*D*) concomitant with accumulation of polyamine metabolites (*E*). While methionine cycle metabolites are enriched overall, individual metabolites associated with glutathione synthesis appear to be decreased, while those responsible for carrying methyl groups and interacting with the folate cycle are increased (*F*). Individual metabolites associated with pyrimidine synthesis are increased in SCD during baseline health (*G*).

### Acute pain events are associated with decreased plasma amino acids in individuals with SCD

Most differentially abundant metabolites in SCD Acute relative to SCD BL were decreased (**Figure 1B, F-H**). Consequently, few pathways were enriched among the increased metabolites. These included pathways associated with phenylacetate and carbohydrate metabolism (**Figure 3A**). Conversely, downregulated metabolites were enriched for a breadth of metabolic pathways, including pathways associated with lipid utilization, antioxidants, and amino acid metabolism (**Figure 3B**). Affected amino acids include serine, asparagine, glutamine, isoleucine, cysteine, proline, and arginine (**Figure 3C**, **S. Table 1**), which were already diminished in SCD at baseline health. While methionine itself was not decreased during acute pain in SCD, metabolites downstream of methionine were significantly decreased (**S. Table 1**). Therefore, while various amino acid metabolites were increased or decreased between SCD baseline health and healthy controls, amino acid metabolism is nearly ubiquitously downregulated during acute pain in SCD.

**Figure 3.**
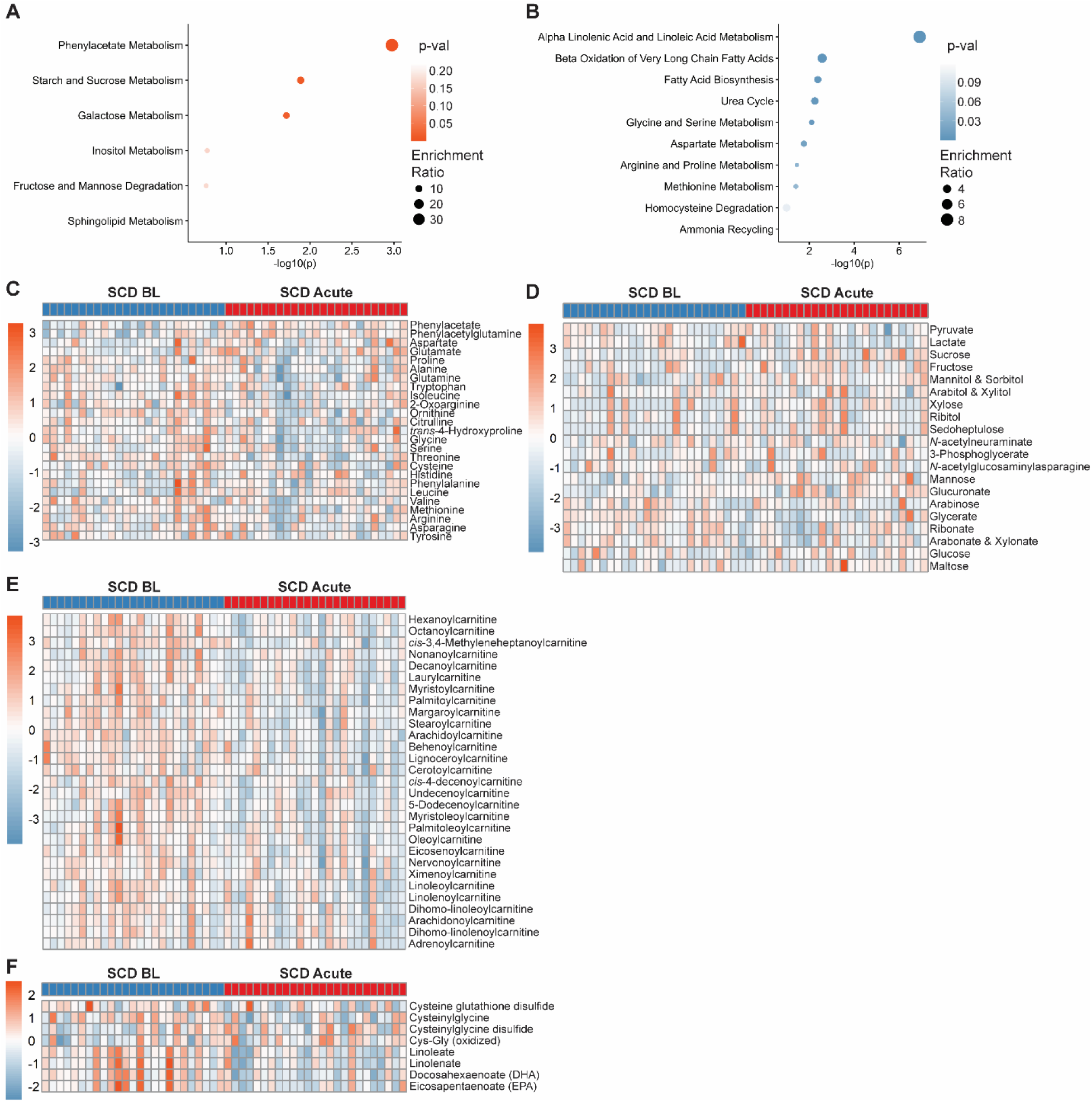
Acute pain in SCD exhibits a metabolic signature consistent with hypoxia. (*A*) MSEA of upregulated metabolites during acute pain in SCD reveals enriched carbohydrate metabolism. (*B*) Metabolic pathways associated with amino acid metabolism, oxidative bioenergetics, and mitigating oxidative stress are downregulated during acute pain in SCD. (*C*) Many amino acids and their metabolites are downregulated in SCD Acute samples compared to SCD BL. (*D*) Despite enrichment of carbohydrate metabolism, few metabolites associated with glycolysis are dysregulated during acute pain. (*E*) Acyl-carnitines are uniformly downregulated during acute pain in SCD, consistent with globally decreased β-oxidation. (*F*) Methionine metabolites associated with glutathione production are decreased in acute SCD pain, as are metabolites in the α-linolenic acid metabolic pathway, indicated reduced antioxidant support.

### Carbohydrate metabolism, β-oxidation, and antioxidant response are altered in individuals with SCD during acute pain

The upregulated and downregulated pathways in the acute pain SCD group were consistent with a shift in bioenergetics. While pathways associated with carbohydrate metabolism were elevated during acute pain (**Figure 3A**), glycolysis metabolites were largely unchanged (**Figure 3D**). Meanwhile, downregulated metabolites were significantly enriched for β-oxidation (**Figure 3B**). Nearly all acyl-carnitines detected in our analysis were significantly decreased in SCD Acute samples (**Figure 3E**), as were ketone bodies, another bioenergetic fuel that relies on oxidative phosphorylation (**S. Table 1**), and components of the tricarboxylic acid cycle (**S. Fig 1**). These findings support a bioenergetic shift consistent with hypoxia. Increased ROS is another common feature of hypoxia. Indeed, we observed a significant reduction of direct ROS scavengers, such as glutathione-related metabolites, and pathways that upregulate ROS scavenging, like synthesis of eicosapentaenoic acid (EPA) and docosahexaenoic acid (DHA) from α-linolenic acid^20^ (**Figure 3F**).

### Network analysis identifies metabolic modules associated with disease state and clinical pain history in individuals with SCD

To determine whether certain metabolites or metabolic pathways were associated with SCD disease state and frequency of pain events in individuals with SCD, we performed weighted correlation network analysis on metabolites of known structure in our dataset. Network analysis revealed eight modules of metabolites, which we then correlated to sample group (**Figure 4A**). The turquoise module had a strong correlation to healthy controls, and the turquoise eigengene value was significantly decreased in healthy controls relative to SCD baseline health and SCD acute pain groups (**Figure 4B**). Meanwhile, the brown and blue modules were strongly correlated with SCD acute pain, and their eigengene values were significantly decreased compared to both healthy controls and baseline health in SCD (**Figure 4C-D**). We identified hub metabolites in these metabolite modules. Consistent with the role of porphyrin metabolism in SCD, hub genes in the turquoise module were enriched for bilirubin degradation products and biliverdin (**Figure 4E**). While a substantial proportion of porphyrin metabolic products were elevated in SCD during baseline health (**S. Fig 2A**), porphyrin metabolites were largely unaltered during acute pain in SCD relative to SCD baseline health. Hub metabolites in the brown module were enriched for free medium-and long-chain fatty acids (**Figure 4F**), and hub metabolites in the blue module were long-chain phosphocholines and phosphoethanolamines (**Figure 4G**). Together, these modules reveal dyslipidemia as a major correlate of acute pain in SCD.

**Figure 4.**
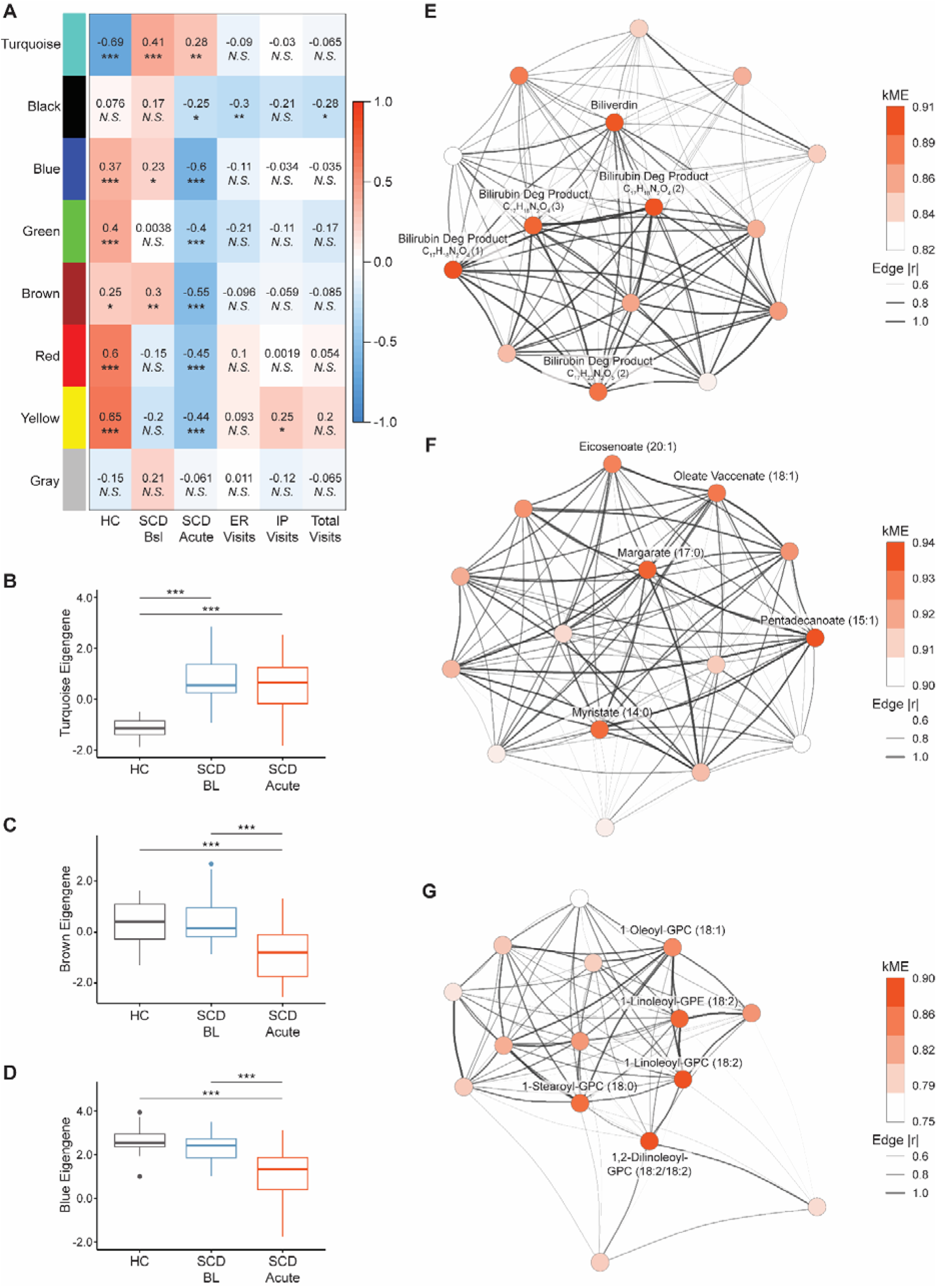
Network analysis identifies metabolite modules associated with baseline health and acute pain in SCD. (*A*) Weighted gene correlation network analysis (WGCNA) performed on known metabolites results in a network with 8 modules, which correlated to group (HC, SCD BL, or SCD Acute) and the number of pain-related emergency room visits (ER visits), inpatient hospitalizations (IP visits), and combined number of ER and inpatient visits (Total visits). (*B*) The turquoise module eigengene is increased in SCD samples regardless of pain presentation, while the brown (*C*) and blue (*D*) modules are significantly reduced in SCD Acute compared to HC and SCD Bsl. (*E*) Visualization of the turquoise module network and hub metabolites reveals enrichment for biliverdin and bilirubin degradation products (Bilirubin Deg Product). (*F*) Hub metabolites within the brown module are enriched for free medium-and long-chain fatty acids. (*G*) Hub metabolites within the blue module are not as densely connected as the turquoise or brown modules, and are enriched for long-chain phosphocholines, lyso-phosphocholines, and lyso-phosphoethanolamines. (A) WGCNA with Pearson’s correlation; *N.S.* indicates p>0.05, * indicates p<0.05, ** indicates p<0.01, *** indicates p<0.001. (*B-D*) One-way ANOVA with Tukey’s *post hoc* test, *** indicates p<0.001. (*E-G*) Network graphs of the top 15 hub metabolites for each module with the top 5 hub metabolites labeled. Nodes are colored according to normalized correlation of each metabolite to the module (kME) and edge thickness indicates strength of connections between metabolites within a module.

To correlate metabolic modules with clinical outcomes in individuals with SCD, we used the rate of acute care visits, defined as the number of emergency room (ER) visits, inpatient hospitalizations (IP) visits, and total acute care visits (*e.g.*, sum of ER and inpatient visits), as measures of acute pain frequency. The only module to significantly correlate with ER visits or total acute care visits was the black module (**Figure 4A**), yet the black module eigengene value was not significantly different between groups (**Figure 5A**) and demonstrated weaker connectivity than other modules (**Figure 5B**). The yellow module, however, was significantly associated with inpatient visits and was significantly decreased in the SCD baseline health group and SCD acute pain group (**Figure 5C**). Hub metabolites within the yellow module were all long-and very-long-chain sphingomyelins (**Figure 5D**). Hub metabolites associated with other modules are presented in supplementary table **S. Table 2**. Together, these findings identify dyslipidemia as a potential driver of acute pain and pain frequency in children and adolescents with SCD.

**Figure 5.**
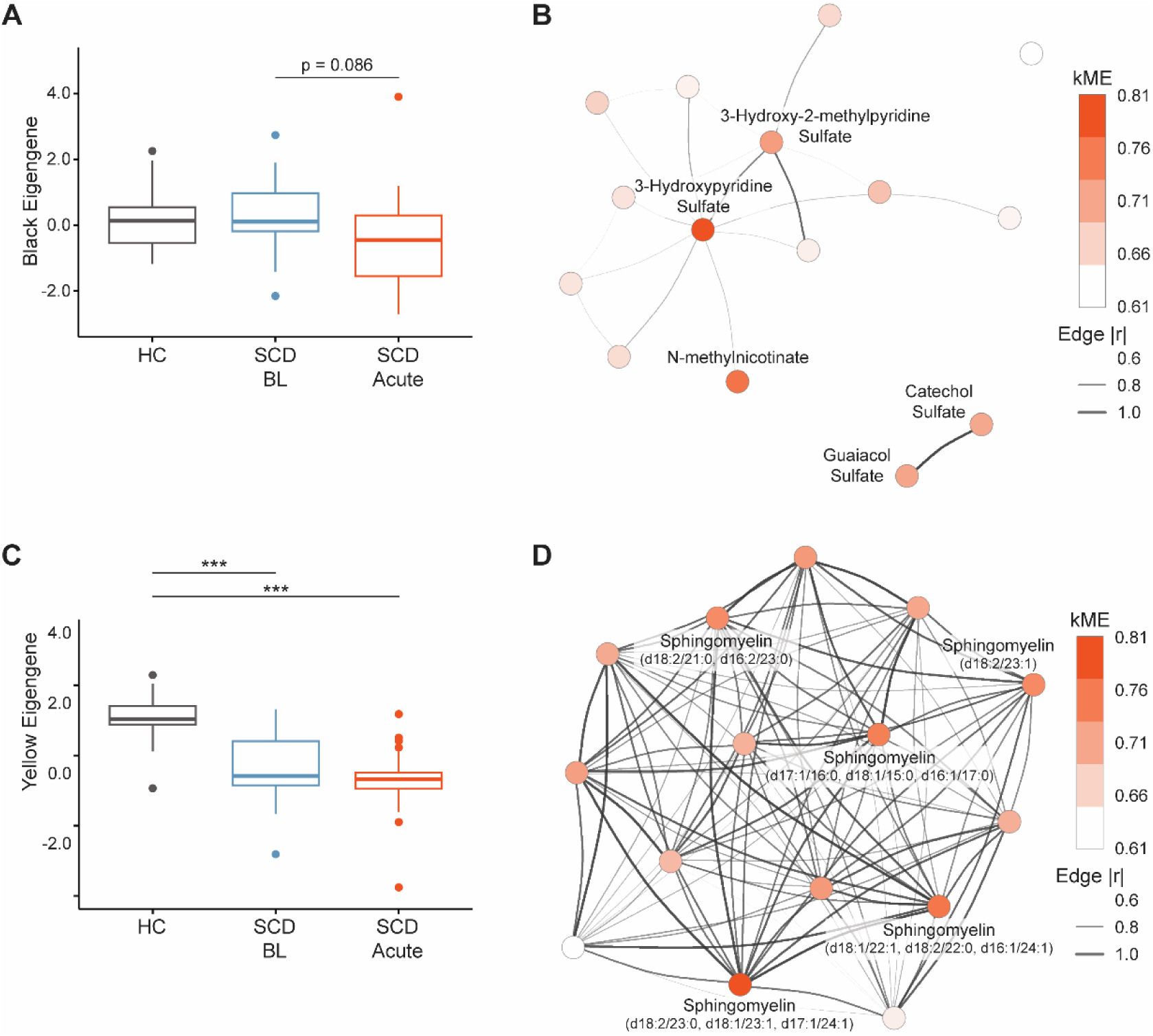
Sphingomyelins are hub metabolites in a metabolic module associated with SCD acute pain. The black metabolite module (*A*) is not significantly altered between healthy controls, SCD baseline health, and SCD acute pain, while the yellow module (*B*) is significantly decreased in SCD baseline health and acute pain. (*C*) Hub metabolites within the black module exhibit weaker connectivity and are associated with pyrimidine synthesis. (*D*) The top hub metabolites in the yellow module are exclusively long-chain sphingomyelins. (*A-B*) One-way ANOVA with Tukey’s *post hoc* test, *** indicates p<0.001. (*C-D*) Network graphs of the top 15 hub metabolites for each module with the top 5 hub metabolites labeled. Nodes are colored according to normalized correlation of each metabolite to the module (kME) and edge thickness indicates strength of connections between metabolites within a module.

**Figure 6.**
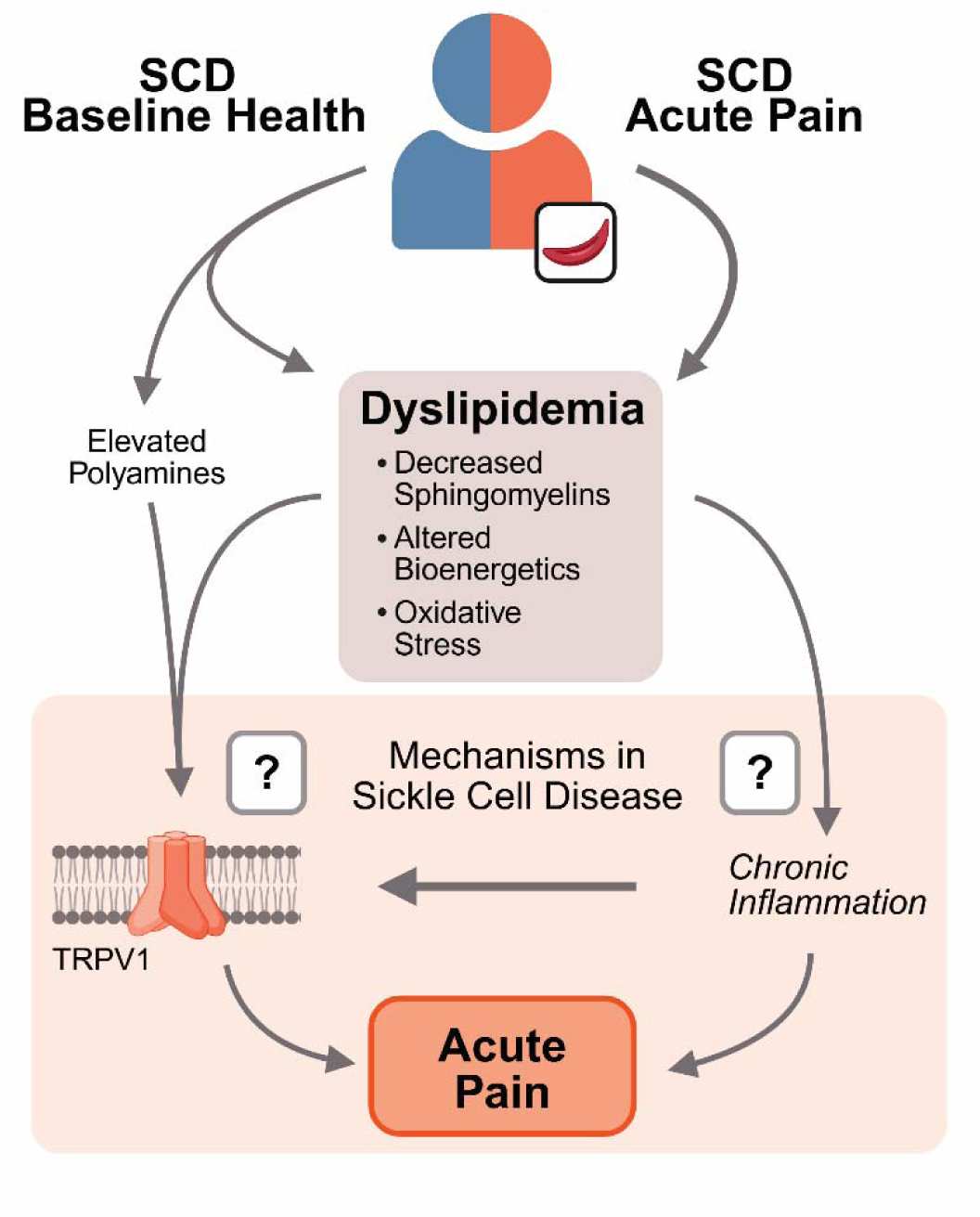
Metabolic signatures of SCD acute pain and baseline health are related to pain neurobiology. Individuals with SCD exhibit elevated circulating polyamines and features of dyslipidemia during baseline health. Circulating lipid profiles are further altered during acute pain in SCD, suggestive of altered bioenergetics and increased oxidative stress. Both elevated polyamines and dyslipidemia have been associated with increased TRPV1 activity and chronic inflammation associated with other etiologies of acute and chronic pain. It remains unknown whether these factors are merely correlative or indeed cause acute pain in individuals with SCD.

## Discussion

Individuals with SCD experience severe episodic acute pain secondary to complex biology partially driven by vaso-occlusive events and acute ischemic and inflammatory injury^21,22^. Despite the lifelong nature of severe and debilitating acute pain among individuals with SCD, the molecular and metabolic features remain poorly understood. We used untargeted metabolomics to profile changes in the plasma metabolome in children and adolescents with SCD during acute pain and during baseline health in the same individuals and compared these to healthy controls. Arginine metabolites were decreased, and methionine metabolites were enriched in SCD during baseline health, leading to increased accumulation of polyamines and decreased metabolites associated with ROS scavenging. Amino acids and their metabolites were further decreased in plasma from individuals with SCD during acute pain relative to baseline health, exacerbating deficiencies in ROS scavenging pathways. Likewise, our data suggest a shift away from oxidative bioenergetics during acute pain in SCD. Finally, we identified metabolic modules and hub metabolites associated with dyslipidemia that strongly correlated with acute pain in SCD and correlated with inpatient hospitalizations for pain. Together, these findings delineate a metabolic fingerprint mirroring systemic hypoxia and indicating a role for dyslipidemia during baseline health and acute pain presentation in children and adolescents with SCD.

Individuals with SCD endure a complex pain experience, characterized by frequent acute pain episodes and the emergence of chronic pain in adolescence. SCD pain is often described as “aching”, “throbbing”, “sharp”, or “burning”, and chronic pain has been described in up to 40% of affected children^21,23,24^. We only detected significantly increased analgesics and analgesic metabolites in plasma from individuals with SCD in the acute pain cohort.

Many metabolites that were elevated or decreased at SCD baseline health were not further elevated or decreased during acute pain. Importantly, this does not preclude their contribution to chronic pain in SCD. For instance, our data show porphyrin metabolism is elevated in pathways and metabolic modules associated with SCD baseline health in our data, and prior work shows it is associated with pain in preclinical models^25^. Hemolysis in SCD leads to elevated cell-free heme in circulation, which is metabolized into biliverdin and subsequently bilirubin. Free heme activates Toll-like receptor 4 (TLR4), sensitizing nociceptors and leading to chronic pain, including pain in SCD^25–31^. Similarly, bilirubin activates transient receptor potential melastatin-containing 2 (TRPM2), directly activating sensory neurons, and antagonism of TRPM2 reverses mechanical hypersensitivity in the Townes model of SCD^32,33^. Polyamine biosynthesis is another pathway enriched during SCD baseline health. Polyamines regulate many cellular processes implicated in pain, including mRNA transport and translation, activation of inflammatory pathways, and regulation of ion channels^34–42^. Indeed, a recent genome-wide association study identified variants associated with the neuronal polyamine transporter, *SLC45A4*, that were associated with chronic pain intensity^43^. Knockout of *SLC45A4* in mice decreased excitability in a subpopulation of nociceptors and reduced response latency to noxious heat. One of the ion channels regulated by polyamines is transient receptor potential channel vanilloid 1 (TRPV1)^40,41^. Prior work from our group and others demonstrated that TRPV1 is sensitized in sensory neurons in preclinical models of SCD, and that TRPV1 antagonism rescues many pain-like behaviors present in these models^44–46^. TRPV1 is also sensitized in induced-pluripotent stem cell-derived sensory neurons from patients with SCD following acute exposure to spermine^47^. Thus, some pathways elevated at baseline health are implicated in chronic pain in SCD.

Consistent with prior reports, we found significant depletion of metabolites associated with arginine metabolism during SCD baseline health, which was exacerbated during acute pain presentation^12,13^. Other amino acids were also decreased during acute pain in SCD, mirroring a metabolic consequence of hypoxia^8,10^. L-glutamine supplementation reduces the frequency of acute pain presentation in patients with SCD^48^, while supplementation with L-arginine has yielded mixed results. A recent meta-analysis suggests that L-arginine supplementation does not affect pain score, though it may reduce time to resolution of acute pain in SCD^49,50^. It is unclear why supplementation with L-glutamine but not L-arginine is protective in SCD. Thus, the effects of these amino acids on various cellular processes, including hypoxia, remain promising therapeutic targets in acute pain during SCD.

Our network analysis identified several metabolic modules associated with lipid metabolism that were significantly decreased during both the SCD baseline health and acute pain groups. Mechanistically, these modules suggest metabolic alterations that support a proinflammatory environment in SCD, leading to pain. The top hub metabolites of one module strongly associated with SCD acute pain were enriched in free fatty acids. At the same time, our data revealed significant downregulation of fatty acid β-oxidation, indicating shifting bioenergetics away from oxidative phosphorylation. Oxidative bioenergetic substrates, such as acyl-carnitines and ketones, were uniformly decreased during acute pain in SCD, supporting altered preference in bioenergetic fuel. The balance of glycolysis and β-oxidation directly affects polarization of macrophage and CD4^+^ T cells, shifting toward proinflammatory populations with reduced mitochondrial oxidative phosphorylation^51–54^. Both macrophage and T cell populations have been implicated in chronic inflammatory pain^55,56^. SCD macrophages have elevated anaerobic glycolysis, reduced mitochondrial mass, and impaired oxidative phosphorylation^57^. Subsequently, SCD macrophages have a stronger inflammatory phenotype. Inflammatory macrophages are directly involved in chronic pain, where the release of cytokines like tumor necrosis factor α (TNFα), interleukin-6, and C-C motif chemokine ligand 2 (CCL2) directly sensitize sensory neurons, contributing to pain^45,58–60^. While the extent to which increased inflammatory macrophage signaling contributes to acute pain in SCD is unclear, transfer of inflammatory macrophages acutely leads to pain-like behaviors in mice, while inflammation-resolving macrophages aid the resolution of inflammatory pain^55^.

Shifting bioenergetic fuel preferences may also directly affect sensory neurons in SCD. Oxidative ketone utilization by mitochondria is required for the normal function of sensory neurons and is antinociceptive in mice and humans^61–64^. Similarly, impaired pyruvate oxidation directly leads to sensitization of sensory neurons and increased pain behaviors in mice^65^. It is currently unknown whether mitochondria are dysfunctional or susceptible to hypoxia in SCD sensory neurons. Thus, future work addressing mitochondrial health in sensory neurons will clarify their contribution to altered bioenergetics during acute pain in SCD.

Only one metabolic module was associated with SCD and correlated with number of inpatient hospitalizations for pain. This module was enriched for sphingomyelins, all of which were decreased in SCD plasma. While others have described decreased sphingomyelins in plasma from individuals with SCD^66^, our study is the first to associate sphingomyelins with pain history, highlighting the potentially important role of sphingolipid metabolism in the pathophysiology of pain in SCD. Although the mechanisms by which sphingomyelins contribute to pain have not been explicitly defined, they likely affect inflammatory pathways and signal transduction. Acid sphingomyelinase activity is increased in sickle RBCs, leading to sphingomyelin breakdown, activation of myeloid-lineage cells, and release of proinflammatory cytokines, like TNFα^67^. Activation of naïve myeloid cells with plasma collected from individuals with SCD likewise led to elevated cytokine release and upregulation of TNFα^22^. Voltage-gated sodium (Na_V_) channels are sensitized by TNFα, as are TRPV1 and other channels, increasing sensory neuronal excitability and contributing to pain^58,60,68–70^. While the role of TRPV1 in SCD is well established^44,45,47^, contributions of Na_V_ channels in SCD pain have not been explored. Activation of the TNFα signaling pathway also increases sphingomyelinase activity^71^, potentially leading to a feedforward loop of dyslipidemia and inflammation. Our results do not address local sphingolipid metabolism in peripheral or nervous tissues on pain in SCD; however, leaving potential pro-or anti-nociceptive roles of sphingomyelins in neuronal tissue unexplored. Sphingomyelin is a key component of lipid rafts, facilitating pro-inflammatory and pro-nociceptive signaling through TLR4 and TRPV1 in sensory neurons^27,72,73^. Depletion of sphingomyelin reverses neuronal sensitization and improves pain behaviors in mice ^72,73^. Signaling downstream of TNFα would hypothetically decrease neuronal sphingomyelins; however, suggesting a nuanced role of sphingomyelins in pain, warranting further mechanistic exploration.

Sphingomyelins are also essential to glial function in the central and peripheral nervous system (CNS and PNS, respectively). Oligodendrocytes in the CNS and Schwann cells in the PNS myelinate axons, providing metabolic and signaling support for nerve function. Sphingomyelins, as the name suggests, are significant components of myelin and are produced by myelinating glia^74^. Aberrant sphingomyelin synthesis and transport directly lead to myelin defects and nervous system dysfunction^75–77^. Similarly, disruption of Schwann cell energetic cooperation with sensory afferents leads to concomitant myelination deficits and reduced sphingomyelin levels^78^. Indeed, we previously reported hypomyelination and myelin defects in the peripheral nerve of Townes SCD mice^79^. Thus, while glial dysfunction has not been explicitly linked to pain in SCD, our current findings linking sphingomyelin deficits to SCD acute pain and suggest glial deficits may contribute to neuropathy in SCD.

Our study identified a unique metabolic fingerprint of children and adolescents SCD during baseline health and acute pain. During baseline health, individuals with SCD exhibited significantly increased circulating polyamines, which were related to arginine deficiency. This arginine deficiency was exacerbated during acute pain in SCD, along with deficits in other amino acids, consistent with a metabolic signature associated with hypoxia. To our knowledge, we are the first to identify metabolic modules associated with lipid dysregulation, especially related to sphingomyelins, correlated to SCD acute pain. This work advances our understanding of potential mechanisms that contribute to pain in SCD. Our work identifies sphingomyelin metabolism as a potential hub for future mechanistic studies and targets for metabolic intervention in acute pain in children and adolescents with SCD.

## Data Availability

Deidentified summary data are available as supplemental data within this manuscript. Deidentified data are available upon reasonable request to the authors.

## Acknowledgements

We would like to thank the wonderful technical support and service provided by Metabolon. We also thank the dedicated research coordinators who collected the samples from individuals and their data. Very importantly, we thank all the children and adolescents that participated in this project and provided our study team with their plasma samples. This work was funded by National Institutes of Health grants R37NS108278 (CLS), R01NS070711 (CLS), K23HL114636-01A1 (AMB), K24HL180995 (AMB), and F32NS138223 (JDE), and support from the Advancing a Healthier Wisconsin Endowment (CLS, ADE, AMB).

## Authorship Contributions

JDE, CLS, AE, and AB conceptualized the experiments and developed the methodology. JDE, VB, and ZK conducted the experiments. JDE analyzed the data. JDE created the original manuscript draft and figure designs. All authors contributed to editing this paper. Supervision was provided by CLS. Funding for the work was acquired by JDE, CLS, AE, and AB.

## Supplemental Figures

**S. Figure 1.**
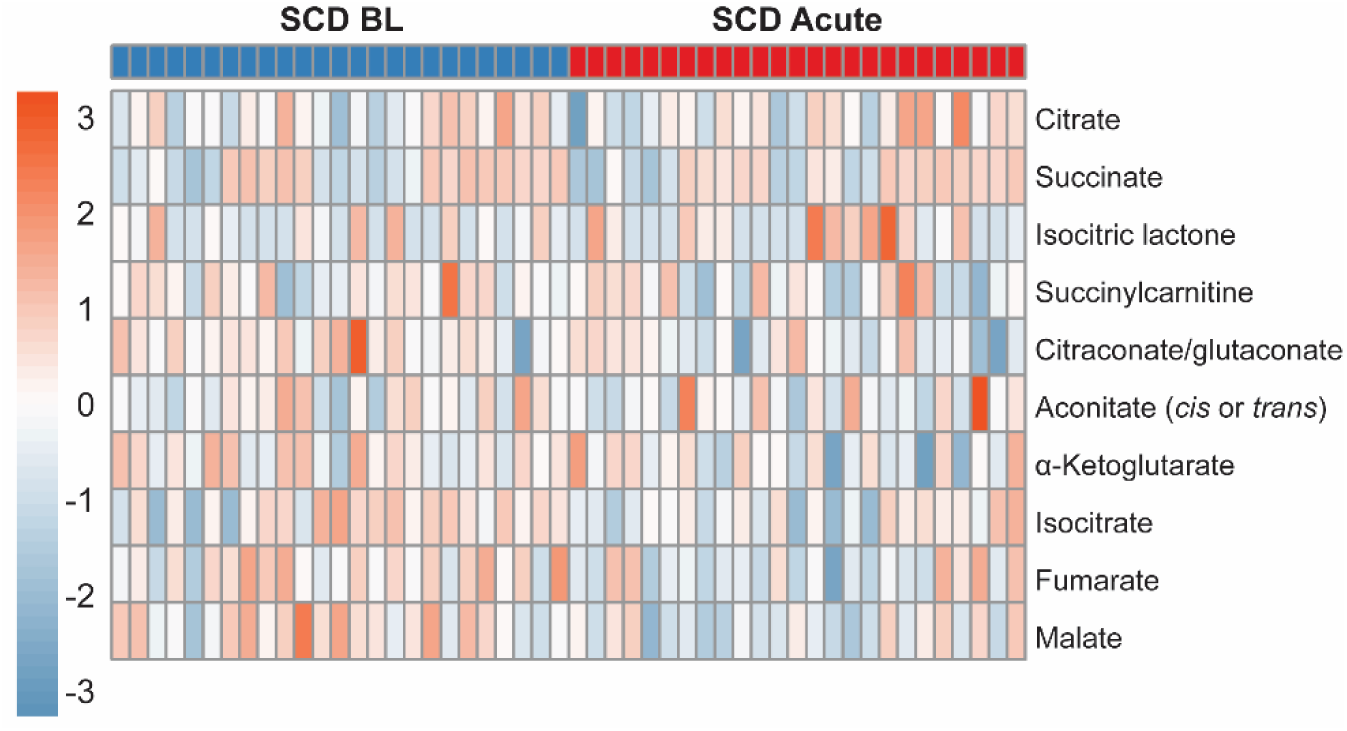
Tricarboxylic acid metabolites are dysregulated during acute pain in SCD. Individual tricarboxylic acid metabolites are decreased in plasma from children and adolescents with SCD during acute pain compared to SCD baseline health.

**S. Figure 2.**
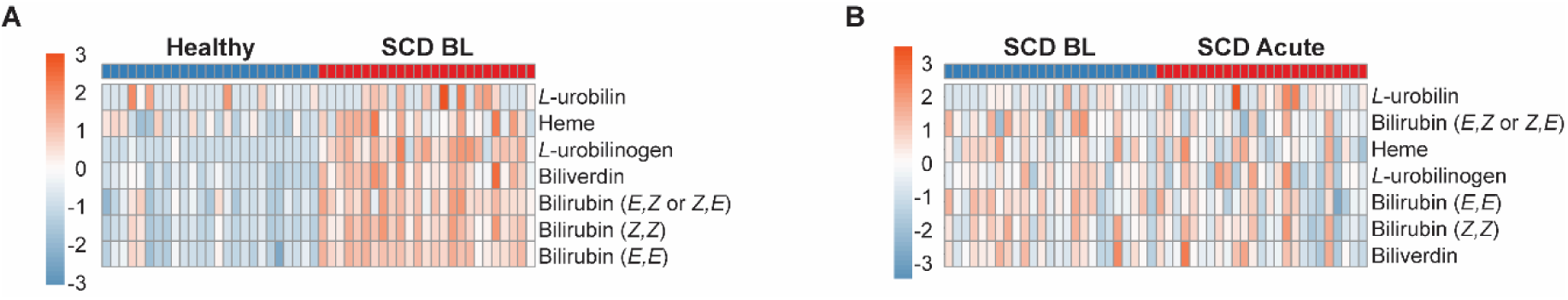
Porphyrin metabolites are increased during baseline health in SCD. (*A*) Metabolites associated with porphyrin metabolism are likewise elevated during baseline health in SCD, but not further elevated during acute pain (*B*).

